# Barriers and facilitators to men’s engagement with digital mental health screening in Estonia: An interpretive qualitative study of user archetypes and design implications

**DOI:** 10.64898/2026.05.12.26353064

**Authors:** Marianne Küüsvek, Riina Hallik, Maarja Pajusalu, Arvi Kuura

## Abstract

Mental health issues are prevalent among men, yet help-seeking remains low due to stigma, masculinity norms, and access barriers. Digital mental health (DMH) screening questionnaires offer opportunities for early detection, but their uptake among men remains limited. This study explored the barriers and facilitators influencing men’s willingness to use DMH screening questionnaires, with the aim of informing user-centered design that supports early detection and engagement. An interpretive qualitative study was conducted through semi-structured interviews with 17 purposively sampled Estonian men aged 20–54 in a highly digitalized context until data saturation was reached. Thematic analysis followed a mixed deductive–inductive approach, combining codes derived from the Technology Acceptance Model, Health Belief Model, User-Centered Design, and Behavioral Design with themes emerging from participants’ responses and evaluations of four screening questionnaires (PHQ-2, PHQ-9, EEK-2, and WHO-5). Key barriers included data privacy fears, distrust of digital solutions, lengthy questionnaires, and poor user experience, while facilitators included anonymity, institutional trust, short mobile-optimized questionnaires, personalized feedback, and clear next steps. As a main contribution, four archetypes were identified: Skeptic, Self-Manager, Explorer, and Situational Seeker, reflecting distinct patterns in privacy concerns, institutional trust, user experience preferences, and help-seeking orientations. Skeptics preferred anonymous, low-friction interactions and demonstrated low institutional trust, whereas Self-Managers valued autonomy, transparency, and evidence-based support. Explorers showed openness toward experimentation and engagement through intuitive and interactive design, while Situational Seekers engaged episodically depending on context and immediate need. Men’s uptake of DMH screening questionnaires is therefore influenced by a combination of social, psychological, and usability factors. Effective design should integrate anonymity, institutional credibility, transparency, user control, and actionable personalized feedback to support engagement and early mental health detection. The proposed archetypes provide a more actionable alternative to demographic and one-size-fits-all approaches for designing DMH questionnaires tailored to male users.

## Introduction

The growing burden of mental health disorders places increasing strain on healthcare systems worldwide, highlighting the urgent need to shift toward earlier, self-directed forms of support [1]. Early self-help and preventive approaches have the potential to alleviate pressure on traditional services and treatment pathways [2]. However, their effectiveness largely depends on individuals’ willingness to acknowledge mental health difficulties and seek help [3,4]. Despite this potential, engagement remains low due to a range of interrelated factors, including limited awareness and mental health literacy, uncertainty about where to seek support, stigma, and culturally embedded social norms [5,6], economic constraints, and issues related to the availability and accessibility of relevant services [7,8]. This challenge is particularly pronounced among men, who are disproportionately affected by depression and anxiety and are less likely to recognize symptoms, perceive a need for care, or engage with conventional mental health services [7,9], often relying on self-management instead [10]. Supporting this, the World Health Organization (WHO) European Region emphasizes that gender norms, social roles, and broader social, economic, and cultural determinants significantly shape men’s health behaviors and their engagement with healthcare services [11,12]. Evidence from Estonian men mirrors international patterns, as a 2023 survey [13] shows that they demonstrate reluctance to seek help, low emotional literacy, and a preference for self-managed solutions, paralleling findings from Europe [8] as well as multinational evidence [14]. Broader research also highlights factors influencing engagement with self-managed and digital mental health approaches [15].

Several initiatives have been implemented to improve men’s access to mental health support, including targeted awareness campaigns and efforts to reduce stigma and promote early help-seeking behaviors [16], as well as workplace mental health programs [17]. While these efforts have shown some positive outcomes, challenges such as low engagement, usability-related burden, and stigma-related barriers [15] continue to limit their impact, highlighting the need for innovative, accessible approaches. Digital solutions, in particular, can overcome geographic, social, and cultural barriers [18], while addressing stigma-related concerns and men’s preference for autonomy in help-seeking [16], which may enhance engagement.

### Digital mental health solutions

Digital mental health (DMH) solutions constitute a heterogeneous group of technologies designed to support mental health assessment, clinical and self-monitoring, and interventions [18,19], encompassing a wide range of modalities beyond any single platform [20]. Empirical evidence indicates that digital mental health interventions are primarily delivered through web-based programs, videoconferencing platforms, smartphone applications, and SMS-based approaches [21]. These approaches provide remote support for mental health conditions affecting nearly one in seven people globally [22]. By improving access and enabling self-monitoring, these technologies complement traditional services with scalable and flexible solutions [4]. Importantly, DMH solutions can help reduce stigma by offering anonymous and self-directed access to support [15,23]. Fully automated and evidence-based DMH solutions, such as digital screening platform integrating questionnaires like the PHQ-9 [24], can facilitate early detection, provide feedback, and help guide users to professional support when needed [25,26], particularly in the context of known barriers to help-seeking [7]. These approaches leverage technological innovation to increase service reach, health literacy, efficiency, and impact [27,28]. Despite advances in digital interventions, practical challenges remain, as many DMH solutions lack clarity, adaptability, while user involvement in their development is frequently limited [29]. Addressing these challenges requires a deeper understanding of underlying factors, which is critical to designing interventions that are not only acceptable but also effective and engaging for men.

### Theoretical frameworks

This study is grounded in multiple complementary theoretical frameworks that collectively inform the understanding of technology acceptance, health behavior, user experience, and behavioral design in the context of digital mental health questionnaires. To understand how DMH screening questionnaires can be effectively adopted and meaningfully integrated into men’s mental health practices, this research draws on several complementary theoretical frameworks, including Technology Acceptance Model (TAM) [30], the Health Belief Model (HBM) [5], user-centered design (UCD) [31], and behavioral design (BD) [32].

Among these frameworks, TAM and HBM serve as the primary explanatory models, addressing two key dimensions of digital mental health (DMH) use: technological adoption and health-related decision-making. TAM explains users’ intention to adopt digital solutions through perceived usefulness and ease of use [30]. These factors are closely linked to usability and functionality in digital mental health platforms [15]. HBM complements this perspective by focusing on how health-related beliefs shape preventive behavior [5]. In men’s mental health, help-seeking is often influenced by perceived stigma and masculine norms, which may act as barriers to recognizing the need for support [23]. Together, TAM and HBM provide a combined framework for understanding both the acceptance of digital solutions and the psychological factors underlying help-seeking. At the same time, UCD and BD translate this perspective into practice: UCD emphasizes designing technologies around users’ needs and experiences [33], while behavioral design supports sustained use and behavior change through targeted design strategies [18].

In addition, theory-informed frameworks can improve adoption, usability, and engagement with DMH questionnaires by addressing men’s psychological readiness, motivation, and contextual factors. Psychological theory can inform the design of interventions that promote user engagement [18], while considering real-world user context is essential for effective digital mental health interventions [27]. Situating this study within broader human-computer interaction (HCI) [34] and service design paradigms further underscores the need for DMH questionnaires to be user-centered, interactive, and aligned with service delivery contexts [19,35].

However, existing frameworks have important limitations, as sociocultural models often treat men as a homogeneous group [36] and do not fully account for how masculinity norms shape help-seeking behaviors [23,37]. Moreover, these frameworks rarely account for how digital environments may reshape such norms [25] by providing anonymous and nonjudgmental spaces [38,39] that can facilitate help-seeking. Psychological models such as HBM [5] and TAM [30] tend to emphasize rational decision-making, while engagement is also shaped by emotional, relational, and contextual factors that are less explicitly captured in these models [14,15]. Design-focused approaches can improve usability and accessibility [40] but do not necessarily translate into sustained engagement or meaningful behavioral change [41,42]. Therefore, a combination of these frameworks is required to provide a more holistic and multi-dimensional understanding of men’s engagement with DMH screening questionnaires.

### Research rationale, aim and questions

Previous research shows that stigma and masculinity norms influence men’s help-seeking behaviors [23,37], shaping attitudes toward emotional expression and support-seeking [16,43]. Engagement with DMH solutions remains consistently low in real-world settings, with studies reporting that only about 4% of users are active on a daily basis [44].

This limitation is particularly relevant in men’s mental health, where stigma and masculinity norms strongly influence help-seeking, emotional disclosure, and perceived acceptability of support [23,37]. Accordingly, it remains insufficiently understood how sociocultural, psychological, and design-related factors interact to shape sustained engagement with DMH screening solutions.

Estonia provides a particularly informative context for this study due to its highly developed digital health infrastructure and strong adoption of e-health services, with public use of online health services well above the EU average across population groups [45]. Consequently, Estonia is conceptualized as a critical case [46], enabling examination of whether engagement barriers persist under conditions of maximal digital accessibility and readiness.

The current study aims to address this gap by examining the factors that shape men’s willingness to use DMH screening questionnaires for early mental health detection. Specifically, it investigates how social norms, masculinity-related help-seeking patterns, and perceived stigma influence engagement with DMH questionnaires [23], as well as how personal preferences and user experience factors shape use and discontinuation [14,15]. Therefore, we target the following research questions:

RQ1: How do social norms, perceived stigma, and individual preferences for digital platforms (e.g., anonymity, accessibility) influence the acceptability and usability of DMH screening questionnaires among men?
RQ2: Which service design and behavioral design approaches best support men’s engagement with DMH screening questionnaires?
RQ3: How do features of the screening questionnaire (e.g., clarity, length, format) influence men’s willingness to use these solutions?

Understanding these factors is essential for developing DMH questionnaires that are more acceptable, engaging, and effective for men, thereby supporting earlier detection and intervention.

## Methods

A qualitative interpretive design [47], grounded in an interpretivist perspective [48], was employed, using individual in-depth semi-structured interviews [49] to explore participants’ experiences with digital mental health questionnaires. Data were analyzed using reflexive thematic analysis [50], resulting in the identification of four participant archetypes. Consistent with prior research using archetypes to categorize recurring behavioral patterns among users [51], these archetypes were developed based on recurring patterns in participants’ experiences and were used as an analytical framework to identify key barriers and facilitators (i.e., user pain points) in help-seeking and engagement with digital mental health questionnaires. In addition, the archetypes informed the development of user experience (UX) principles and guided the selection and evaluation of screening questionnaires.

### Data Collection

Data was collected through individual semi-structured interviews to obtain in-depth accounts of participants’ experiences, perceptions, and opinions [52]. The interviews were conducted using a semi-structured interview guide exploring participants’ experiences, challenges, and needs related to mental health services, with particular attention to the design, usability, and perceived suitability of digital mental health questionnaires for men. The full interview guide, which was reviewed and validated by two external advisors, is available as Appendix 1.

During the interviews, participants were shown four mental health screening instruments: PHQ-2 [53], PHQ-9 [24], EEK-2 [54], and WHO-5 [55]. Participants were asked to review and provide feedback on each instrument, focusing on content, clarity, usability, and suitability. The EEK-2 is widely used in Estonian clinical practice, PHQ-9 in research settings, the WHO-5 was included as a comparative benchmark for phrasing and response format, and PHQ-2 was included for its brevity and established validity as an ultra-brief pre-screening questionnaire.

Seventeen male participants aged 20–54 years were recruited using purposive sampling based on predefined inclusion criteria (male gender and age between 20 and 54 years). Participants were selected to represent diverse life and occupational contexts, including students, managers, entrepreneurs, military personnel, employees, and athletes, in order to capture a broad range of perspectives [56]. Recruitment was intentionally balanced across different age groups (20–29, n=5; 30–39, n=6; and 40–54, n=6) to ensure diversity of perspectives (see Table 1). Snowball sampling was used as a supplementary strategy to facilitate recruitment in age groups where access to participants was more limited.

**Table 1.** Participant characteristics by archetype.

| Archetype | n | Participant (age group) |
| --- | --- | --- |
| **Self-manager** | 3 | P6 (20–29); P3 (30–39); P16 (40–54) |
| **Explorer-type user** | 6 | P1 (20–29); P4 (20–29); P8 (20–29); P9 (30–39); P15 (40–54); P17 (40–54) |
| **Situational seeker** | 3 | P7 (20–29); P11 (30–39); P13 (40–54) |
| **Skeptic** | 5 | P2 (30–39); P10 (30–39); P12 (30–39); P5 (40–54); P14 (40–54) |

Interviews were conducted face-to-face or via Microsoft Teams, according to participant preference. The average interview duration was 29 minutes. Data saturation was assessed iteratively throughout data collection and analysis and was reached at approximately the 15th interview, when no new themes emerged, consistent with established approaches to saturation in qualitative research [57]. Two additional interviews were conducted to improve representation of the oldest age group (40–54); these did not yield new insights and thus confirmed that saturation had been achieved.

Participants received an information sheet in advance outlining the study purpose, procedures, data handling, and their rights, including voluntary participation and the right to withdraw at any time. Verbal informed consent was obtained prior to the interviews, including consent for audio recording, pseudonymization, and transcription. At the beginning of each interview, participants confirmed their understanding and provided verbal consent, which was audio recorded. Verbal consent was considered appropriate given the minimal-risk nature of the study and the use of remote interviewing. As the study did not involve the collection or analysis of participants’ personal health data, formal ethics committee approval was not required under applicable guidelines. Participants were instructed not to disclose sensitive personal health information. The researcher introduced herself at the start of each interview and clarified that she is not a healthcare professional. She explained that the study aims to understand participants’ experiences, opinions, preferences, and attitudes toward digital questionnaires for early mental health detection, in line with principles of reflexivity [58].

All data were pseudonymized. Audio recordings were transcribed using a password-protected web-based transcription service provided by Tal Tech’s Laboratory of Language Technology (https://tekstiks.ee), with access restricted solely to the researcher. Both the audio recordings and the resulting transcriptions were deleted after transcription. Access to the data was restricted to the primary researcher. Data collection and storage complied with the General Data Protection Regulation (GDPR) [59] and the Estonian Personal Data Protection Act [60].

### Data Analysis

Data were analyzed using a combination of deductive coding informed by theoretical frameworks [50,61] and inductive coding that captured insights emerging directly from participants’ accounts [62]. This allowed for the mapping of men’s attitudes towards DMH screening questionnaires.

To ensure familiarization with the data, all interview transcripts from the 17 participants were read repeatedly, and detailed notes were taken to capture preliminary observations and potential coding segments [50]. A systematic manual coding process was conducted using Figma to organize and refine codes. Deductive codes were derived from the study’s conceptual focus areas, including masculinity norms, stigma, privacy and anonymity, usability and design preferences, questionnaire clarity, and motivation to engage. Inductive codes were developed to capture novel insights emerging from the data, such as unexpected barriers, context-specific preferences, and situational nuances. To strengthen analytical rigor, preliminary codes and emerging thematic structures were discussed among the authors [63].

Codes were iteratively grouped into categories and subcategories and subsequently organized into higher-order themes using axial coding [64], focusing on identifying relationships between categories and integrating them into higher-order themes [65]. Themes were repeatedly reviewed and refined and were merged or split when necessary to ensure they accurately reflected participants’ perspectives. Subcategories captured more detailed aspects of participants’ experiences, preferences, and challenges. The final themes were interpreted within the study’s theoretical framework to link the findings with existing theories. The primary themes were developed through a combined deductive-inductive approach and were subsequently organized in relation to the research questions (RQ1–RQ3).

Findings were organized into key themes and participant archetypes, based on shared patterns in participants’ views and experiences, allowing for a clear comparison of perspectives. These archetypes are not intended to represent fixed personality types, but rather flexible patterns that describe how attitudes and behaviors may shift depending on context and conditions. Participants may move between archetypes over time as their circumstances or experiences change, highlighting that these patterns are dynamic, context-dependent, and not fixed [66].

## Results

Based on the responses of 17 participants, four archetypes were identified (see Table 2). The diversity of the sample enabled an exploration of attitudes towards digital mental health screening questionnaires across these archetypes.

**Table 2.** Participant archetypes.

| Archetype | Key characteristics | Representative quote |
| --- | --- | --- |
| **Skeptic** | Distrusts digital solutions; prefers traditional channels; concerned about security and data privacy | *“In fact, these digital questionnaires don’t really provide concrete, strong feedback. You don’t really perceive that they would be of any help.” (P14)* |
| **Self-manager** | Identifies own needs; plans and uses digital solutions deliberately; seeks evidence-based support | *“But actually, it could be something ongoing, to monitor even when everything is fine.” (P3)* |
| **Explorer** | Curious and experimental; searches for the best and most suitable support; takes risks in digital environments | *“If there were a free trial to try it out, I would definitely test the digital version first.” (P4)* |
| **Situational seeker** | Uses digital solutions only in emergencies; engages briefly; seeks quick support | *“Coming from a masculine environment, military/police, well, you don’t go to the doctor unless you’re forced to.” (P13)* |

Table 3 presents a comparative overview of the four participant archetypes across key dimensions, including privacy and data use, institutional trust, preferred user experience, accessibility needs, and help-seeking orientation. The table highlights how each archetype differs in its attitudes, expectations, and engagement patterns with digital mental health solutions. This structured comparison supports a clearer understanding of how user needs and corresponding design requirements vary between archetypes. A more detailed and comprehensive version of this comparison is provided in Appendix 2.

**Table 3.** Comparison of Participant Archetypes Across Key Dimensions.

| **Aspect** | **Skeptic** | **Self-Manager** | **Explorer** | **Situational Seeker** |
| --- | --- | --- | --- | --- |
| **Privacy and data use** | high concern about data misuse; prefers anonymity | wants transparency and control over personal data | Context-dependent: accepts data use when the value is clear. | Context-dependent: willing to share if the need is immediate. |
| **Institutional trust** | low trust in institutions and formal healthcare. | prefers credible, expert-backed support | more open to familiar and trustworthy platforms | Context-dependent: trust depends on the situation. |
| **Preferred UX** | needs a simple, low-friction interface | values a structured and efficient flow | responds well to interactive and visually clear design | needs fast, mobile-friendly, and straightforward use |
| **Accessibility needs** | low-threshold access is essential | flexible access fits daily routines | convenience and low-cost support engagement | immediate availability is important |
| **Help-seeking orientation** | tends to delay help-seeking and rely on self-reliance | uses self-monitoring and seeks support purposefully | open to support when it feels relevant. | Context-dependent: help-seeking is triggered by a specific need. |

facilitators
context-dependent
barriers

As illustrated in Table 3, the comparison does not identify a single dominant archetype. Instead, it highlights substantial variation across users, underscoring the complexity and multidimensionality of help-seeking behaviors in this context.

For men, barriers to help-seeking or to identifying the need for support emerged first and foremost at the level of self-recognition. Participants described difficulties in noticing and acknowledging their own needs: “*Monitoring oneself is relatively difficult, and others tend to notice changes*” (P13), as well as the low prioritization of mental health in comparison to other health concerns: “*Addressing mental health competes with other health needs, for example visiting the dentist*” (P14). A preference for self-reliance: “*Men do not seek help themselves*” (P15) and preconceptions regarding the inaccessibility of services: “*The service is inaccessible, and one would hardly even consider joining a waiting list*” (P10) further impeded help-seeking.

The latter perception was reinforced by awareness of long waiting times for psychological services: “*I am aware that the waiting lists are long; that is a natural part of it*” (P14), the high cost of private consultations: “*The price of a private consultation is 100 euros or more*” (P10), and limited knowledge about available support options: “*People often do not know what options actually exist*” (P15). Interviewees also reported distrust toward general practitioners: “*General practitioners cannot be trusted at all*” (P12) and previous negative experiences with specialists: “*Filling out a mental health questionnaire at the occupational health physician’s office is the most pointless thing ever; it has no meaning whatsoever*” (P16).

Help-seeking was further hindered by fears of potential judgment and occupational consequences, particularly among participants working in security-related professions. This concern was illustrated by one participant (a firearms license holder), who described perceived workplace pressure: “*There is pressure at work about whether you are even allowed to seek help*” (P8). Consequently, participants described a preference for informal support, such as friends and partners: “*Every week my partner and I do these check-ins with each other, like how are you feeling, how are things going*?” (P11). When formal support was considered, personal contact was often valued over digital solutions: “*Personally, I am not really a supporter of those digital solutions; I would prefer a person. I am willing to pay if I can have that person*” (P12).

At the same time, the findings indicate that help-seeking among men appears to be increasingly normalized, with the traditional “be strong” norm gradually receding: *“Men are increasingly willing to seek help, and by 2026, this is already considered the new normal.”* (P5). Stigma was perceived as operating primarily at the societal level rather than within participants’ immediate social environments: “*Perhaps societally it has remained somewhat the case that men do not seek help. Personally, if I do have a problem, I do seek help*” (P1).

The analysis revealed that barriers cannot be strictly separated into technological, social, and regulatory categories, as they are interrelated in digital help-seeking contexts. As one participant emphasized: “*When we are talking about health data or personal matters, it has to be very clear what [data], where, to whom, and for what purpose*” (P13).

Anonymity was considered an important factor by nearly half of the participants, although for different reasons. Some emphasized the need to protect themselves from potential consequences of judgments or diagnoses: “*Since information security is of critical importance, I would prefer anonymous use*” (P6) and “*I need to be sure that no one would use it against me, but rather to help me. That sense of certainty must somehow be built into the system*” (P13). Others were primarily concerned about potential data breaches: “*There is some probability that the data could become public to some extent*” (P3). These findings highlight that concerns about anonymity are shaped both by perceived risks of personal consequences and by fears of data exposure.

Although anonymity was critical for many participants, discussions of personalization revealed ambivalence. Some participants were willing to share personal data if this resulted in clear benefits or improved service quality. On the one hand, personalization was viewed as a means of providing more precise and individualized support: “…*because it can specifically synthesize your problems and concerns*” (P4). Participants also indicated readiness to share data if tangible gains were evident: “*I can share data if I get a better outcome, better recommendations*” (P3). For some, anonymity was not personally important: “*As far as I am concerned, my name can be public; I am not afraid of that. Whether I am anonymous or not*” (P11). However, participants emphasized that personalized solutions would only be acceptable under conditions of transparency and robust data security: “*Personalization does not frighten me if it is clear where the data go and with whom they can be shared*” (P17). Personalization was also associated with the risk of misuse, particularly in relation to diagnostic labeling: “*There is the risk that I receive a diagnosis and that it is misused. Instead of receiving help, I receive a diagnosis*” (P13).

Participants did not necessarily perceive the digital environment as reducing stigma; rather, they expressed additional concerns regarding who has access to personal data and how such data are used. Consequently, anonymity emerged as a facilitating condition under which participants were willing to consider digital solutions: “*If it is sufficiently ensured that confidentiality and anonymity are guaranteed, then I would use it; perhaps a digital solution could even be the first step before counselling*” (P5).

Consequently, anonymity emerged as a facilitating condition under which participants were willing to consider digital solutions: “*If it is sufficiently ensured that confidentiality and anonymity are guaranteed, then I would use it; perhaps a digital solution could even be the first step before counselling*” (P5).

Issues of trust also extended to platform selection. Independent mobile applications were not perceived as sufficiently credible, and commercially oriented web platforms were viewed critically. As one participant explained: “*I think it would have credibility for me if I knew that it was not just an app developed by someone trying to educate and manage me, but that behind it there is, for example, some kind of national healthcare system or a professional association of psychologists*” (P14). Participants expressed a preference for solutions that were publicly administered and institutionally legitimized: “*It should be connected to the national health system*” (P2), as this was associated with greater security and trustworthiness: “*The Health Portal seems logical. It could indeed be centralized. It is sufficiently reliable*” (P12).

Taken together, the interviews revealed that men’s help-seeking is shaped by a complex interplay of personal, social, and technological factors, highlighting the importance of trust, anonymity, and institutionally or publicly administered, accessible, and tailored support for effective engagement.

### User Experience of a Digital Screening Questionnaire

When using a digital mental health questionnaire, poor user experience (UX) emerged as a barrier, often leading to immediate disengagement: *“If the beginning is too complicated, even if it could be very good, I already hit cancel”* (P12). Interviewees emphasized that the application must be simple, clear, and understandable: *“It should be structured in a way that I know where everything is. I shouldn’t have to play detective there”* (P1). Participants also highlighted that collecting data without a clear purpose makes them cautious about using the application: *“If I don’t understand what something is for, I’m more likely not to use it”* (P13).

Regarding user experience, participants primarily valued solutions designed according to mobile-first principles: *“It should be structured for the mobile user interface”* (P3), as well as visual balance: *“It needs to be calm and simple, starting from when I download it”* (P5). Based on their prior experiences, participants preferred a minimalist and neutral visual style, which they believed would help reduce distractions: *“Minimize all that noise, just give me what I need”* (P2). They also emphasized that the application must integrate smoothly into daily life to ensure sustainable use: *“If I were seeking mental health support, it has to integrate into my life, my schedule, and my financial situation. If integration isn’t easy, it probably won’t stay in my life for long”* (P6). In addition, initial guidance supported the use of the digital questionnaire: *“The first time, there could be a walkthrough that shows you every click, guiding you”* (P3). However, some participants preferred to skip guidance: *“I don’t want guidance and skip that part. It should be so simple to use that you can easily understand what needs to be done”* (P9).

Notifications in the digital application were an important component of UX, but their design influenced the user experience in different ways. Interviewees noted that aggressive or overly frequent notifications could provoke negative reactions and lead to discontinuation of use: *“If it starts notifying me too much and disturbing me, I’ll stop using it”* (P13). Conversely, well-timed notifications, for example before going to bed, were considered appropriate: *“The notification needs to be sent at the right moment”* (P10), and notifications under the user’s control were seen as effective and supportive of continued use: *“If there is no notification, I won’t remember either”* (P8). Most participants kept notifications turned off to reduce digital noise: *“I generally have notifications turned off because they cause anxiety”* (P15). However, some preferred receiving an SMS instead of a standard notification when needed: *“I’d rather have an SMS. It’s not deleted so easily and stays noticeable”* (P15).

Finally, but critically, an essential UX element was the feedback provided after completing the questionnaire. Interviewees emphasized that feedback is a key motivator for using a digital screening questionnaire, as it must offer immediate and practical benefits: *“If I know that I actually get something from this feedback”* (P4). Participants expected personalized results that could be tracked over time to observe changes and trends: *“It would be good to be able to monitor your answers and overall health over a longer period to notice trends”* (P6). They also desired concrete guidance on next steps: *“It could provide some instructions for further action. For example, if my case is already serious, I should see a psychologist”* (P5).

In addition to specific instructions, participants wanted the option to share feedback with professionals: *“I would like this information to somehow reach a specialist”* (P14), as well as control over their own data: *“There should be a choice whether to save the results or not”* (P1). In the context of men’s mental health, clarity regarding next steps and the handling of data was particularly emphasized: *“You should definitely get feedback on what happens with the data and what I should do next”* (P13). Without substantive feedback, the digital questionnaire appeared useless to men, often resulting in disengagement.

To conclude, the interviews revealed that men’s engagement with a digital mental health screening questionnaire depends on a clear, simple, and mobile-friendly user experience, seamless integration into daily life, well-timed and user-controlled notifications, and, most importantly, personalized, actionable feedback with transparency and control over data. Poor UX or lack of meaningful feedback often led to disengagement.

### Features of the Digital Screening Questionnaire

Participants described the structural and content-related features of the digital screening questionnaire that shape its perceived suitability and trustworthiness. These features directly influenced their willingness to respond honestly and to complete the questionnaire. For the interviewees, the length of the questionnaire was a crucial factor, with an acceptable upper limit of around 10 items: *“If it gets brutally long, interest disappears. Maximum 10 questions”* (P8). Additionally, the time required to complete it should not exceed 5-10 minutes: *“If the questionnaire takes longer than five minutes, I wouldn’t bother”* (P12). This was primarily justified by concerns about loss of focus and time demands: *“It’s related to the time required. I can only concentrate for a few minutes. A shorter questionnaire would help me complete it”* (P5).

Participants preferred questions to be presented one at a time without the need to scroll, as this approach allows focused attention on each question and reduces both visual and cognitive load: *“It’s uncomfortable when all questions are presented at once; you have to scroll and cannot process each question individually”* (P3). It was also considered important that, after submitting a response, it is clearly indicated as saved and the user proceeds to the next step, supporting a sense of control and understanding of the process: *“The most reasonable way is to have one question per page, then you press it, it gets saved, and you move forward”* (P4). This approach minimized the discomfort caused by displaying all questions simultaneously: “*I would prefer it if there was one question at a time”* (P14). Usability was further supported by the ability to save responses during the questionnaire: *“Saving is very important. If something breaks and I have to start over, it’s frustrating”* (P15), as well as by clearly visible information at the start regarding questionnaire length and progress: *“I want to know the length immediately; if it’s too long, I will quit right away”* (P5).

Regarding the questions in the digital screening questionnaire, participants emphasized the importance of simple, clear, secure, and understandable language: *“There is too much technical terminology. It seems that each platform and person has their own terminology, which is hard to understand”* (P6). They also stressed that questions should be as short and understandable as possible: *“Questions should definitely be short and understandable. I think this is one of the most important components in developing the whole solution. Better to have more questions that are short than long questions”* (P5).

The ability to skip uncomfortable questions was also considered important: *“There should definitely be uncomfortable questions; if a question bothers me, there should be an option to skip it”* (P13). At the same time participants also valued questions that encouraged them to reflect on their own experiences: *“These questions should be such that I actually have to focus and think about my own situations”* (P1). Some interviewees preferred multiple-choice responses: *“I like multiple-choice answers”* (P12), while others highlighted the importance of a free-text field for additional clarification: *“If possible, there should be a place to add a description”* (P13).

In terms of response format, some participants preferred answering on an even-numbered scale (0–5), as this format eliminates a neutral midpoint and encourages the respondent to lean toward one side or the other: *“It’s more correct to ask on an even-numbered scale rather than an odd one, because then I have to take a position, whether I lean one way or the other”* (P3). These interviewees explained that the neutral option often felt like avoiding a decision, whereas an even-numbered scale prompted them to reflect on their true opinion and commit to a position.

Several participants emphasized that the content and relevance of the questionnaire outweigh the importance of its design: *“For me, it doesn’t matter what form the questionnaire takes; the content is more important. And whether I actually believe that the whole thing will be useful to me in the end”* (P10). Although content was considered a priority, participants also highlighted the importance of result design, preferring a combination of text and visuals (e.g., graphs, animations), as this was easier to understand than text alone: *“A combination. For example, scales accompanied by a short explanatory text”* (P15). Several participants specifically noted that combining text with visuals made the results clearer and more accessible.

To sum up, participants described the structural and content-related features of the digital screening questionnaire that shape its perceived suitability and reliability (see Table 4).

**Table 4.** User Experience principles from semi-structured interviews.

| Include/Use | Avoid |
| --- | --- |
| • Maximum of 10 questions<br>• Optimal duration 5–10 minutes<br>• One question visible at a time<br>• Question progress bar visible<br>• Automatic saving of responses<br>• “Skip and answer later” button for uncomfortable questions<br>• Combined multiple-choice answers with an optional free-text field<br>• Use even-numbered scales; final design should be based on both user preferences and literature<br>• Present final results as a combination of short text and visuals (graphs + animations) | • Scrolling through all questions<br>• Avoid complex terminology<br>• Avoid text-only results<br>• Do not use long, rambling sentences: questions should be short<br>• Non-informative or “empty” questions |

Taken together, these findings suggest that user experience in digital screening questionnaires is not determined by design elements alone, but by the interplay between usability and perceived relevance. While streamlined structure and interaction design reduce cognitive load, participants’ willingness to engage ultimately depends on whether the content feels meaningful and beneficial. This underscores the importance of aligning technical design decisions with user expectations and needs.

### User feedback on Selected Screening Questionnaires

In addition to general features, participants also evaluated specific existing screening questionnaires. In their assessments of PHQ-2, PHQ-9, EEK-2, and WHO-5, no clear preference for any single instrument emerged. However, several recurring evaluation criteria were highlighted, including questionnaire length, structural clarity, content depth, and response-related burden.

The PHQ-9 received the most frequent positive feedback for its moderate length, detailed content, and clear structure: *“I would prefer the PHQ-9; it was detailed and not too long” (P4), “PHQ-9, because it seems the easiest”* (P2). The PHQ-2 was praised for its simplicity and visual clarity: *“PHQ-2 has the simplest and clearest structure”* (P15), although some participants wondered if it provided enough information: *“With PHQ-2, I don’t know how much these two questions would really give me”* (P1).

Participants found the WHO-5 clear and of moderate length, especially because the results were easy to understand: *“I would prefer WHO-5. There aren’t too many questions, and the results are clearly explained at the bottom”* (P9), though it was sometimes perceived as too general or brief: *“WHO-5 seems useful, but it’s too short and general”* (P6). The EEK-2 was regarded as professional: *“If I were in greater distress, I would choose the EEK-2. I would pick it because it is considerably more professional”* (P5), but it was seen as too long for daily use: *“EEK-2 is too long. Probably useful, but impossible to complete every day”* (P6). Overall, participants sought a balance between brevity and substantive informativeness, preferring questionnaires with a clear structure and an understandable response format: *“Honestly, I can’t say I’m impressed by any of them. We live in a world that wants quick answers and fast solutions. All of these questionnaires feel like attempts to get a grasp on the matter”* (P16). Participants emphasized the importance of questionnaires that combine clear structure, manageable length, and sufficient detail to allow comfortable responses and easy understanding of the results.

The responses pointed out that users prefer screening questionnaires that combine manageable length with adequate informational depth, clear structure, and easily understandable results.

## Discussion

This study shows that men’s use of DMH screening questionnaires is not determined only by whether the technology is available. It is also influenced by social norms and psychological factors, including masculinity-related barriers to help-seeking behaviors [23,67]. Previous research has identified key barriers like stigma and usability issues in digital mental health solutions [6,68], but the findings show that these factors often occur together and jointly influence how people engage with them. Importantly, no consistent age-related differences were found in this study, consistent with previous evidence suggesting that demographic differences, including age, are not consistently associated with engagement [26].

Building on these findings, this study makes three key contributions. First, it identifies four user archetypes describing how men engage with digital mental health screening solutions. Second, it shows that engagement is context dependent rather than stable, extending traditional behavioral models. Third, it highlights institutional trust as a key determinant of willingness to use digital mental health screening solutions.

These contributions are reflected in the identification of four user archetypes: Skeptic, Self-Manager, Explorer, and Situational Seeker- which reflect different ways men relate to trust, privacy, effort, and help-seeking. Rather than grouping users by demographics, this typology helps explain how and under what conditions men engage with DMH screening questionnaires. Importantly, these archetypes are not fixed or mutually exclusive categories, instead, they represent flexible and context-dependent patterns of behavior, in line with a reflexive thematic analysis approach. Beyond describing user differences, the typology may also have practical value: it helps identify user groups that are more relevant to target with specific engagement strategies and those that may require more tailored and supportive design approaches.

The findings support earlier models but also add new insights. As suggested by the TAM [30], men were more likely to engage when the questionnaire felt useful and easy to use, particularly in relation to its length, clarity, and feedback quality. However, these preferences were not stable. What feels useful can change depending on the situation, particularly for Situational Seekers and Explorers. In a similar way, HBM [5] explains help seeking through perceived need and barriers, but our findings suggest that these are also dynamic and context dependent, shaped by emotional state and social context. Overall, engagement with DMH screening solutions is not fixed but evolves over time through interacting personal, social, and technological factors.

Masculinity norms and stigma continue to influence help-seeking, including in digital contexts [23]. Digital solutions are often assumed to reduce stigma by enabling anonymity [38], but our findings suggest that anonymity alone is insufficient. Participants valued anonymity only when it was combined with trust, perceived safety, and control over personal data. Concerns about data misuse, distrust in institutions, and possible personal consequences were especially common among Skeptics, reflecting broader concerns about privacy and trust in digital mental health [39]. This indicates that digital solutions alone cannot overcome sociocultural barriers. Instead, DMH solutions must address both technical dimensions such as data protection and relational dimensions such as trust and credibility. Preferences for institutionally backed solutions further highlight the importance of perceived legitimacy in engagement [69].

From a practical perspective, the archetypes offer clear guidance for designing DMH screening questionnaires. Different user types need different engagement strategies. Skeptics are unlikely to engage unless there are strong guarantees of anonymity, transparency, and institutional credibility, highlighting the importance of trust and privacy considerations in digital mental health engagement. Self-Managers, in contrast, show higher readiness to engage. They benefit from structured, evidence-based solutions, as well as features such as progress tracking and personalized feedback, which support ongoing self-monitoring. Explorers are more likely to engage with interactive and well-designed solutions, where usability and user experience are central drivers of adoption. Situational Seekers engage only occasionally, typically in response to a specific need.

These findings show that a one-size-fits-all approach is unlikely to work. Instead, design strategies that take different user types into account, or adapt to users’ needs, are more likely to support engagement [18]. The study also has value across several fields. For healthcare professionals, the typology offers a more detailed understanding of how men seek help, which can support more tailored communication and interventions. For designers and developers, the findings point to clear design needs, such as flexible levels of anonymity, simple and structured user flows, and meaningful feedback. For policymakers and service designers, the results highlight the importance of institutional trust and system-level integration in increasing both the credibility and use of DMH solutions.

At the same time, the findings show that designing effective DMH screening questionnaires is complex, consistent with established user-centered design principles [31]. Key factors such as anonymity, personalization, trust, and usability often involve trade-offs rather than simple solutions. For example, increasing personalization usually requires collecting more user data, which can conflict with the need for privacy. Similarly, making questionnaires shorter and simpler may reduce effort for users but can also limit their clinical depth. This means that design decisions cannot focus on just one goal at a time. Instead, they require balancing different user needs and situational factors. As shown in Table 4, engagement depends on how these trade-offs are managed, rather than fully resolved. This highlights the importance of flexible and context-sensitive design approaches [18].

Finally, the findings highlight the need to combine clinical validity with user-centered design. Evidence-based content is important to ensure that screening solutions are reliable and useful, but this alone is not enough. In practice, engagement depends on whether the solution feels easy to use and relevant to the user. Features such as personalized and actionable feedback, clear guidance on next steps, and the ability to track changes over time were especially important for sustained engagement. Without these, even clinically valid solutions may be seen as irrelevant or too demanding.

Overall, this study shows that men’s engagement with DMH screening questionnaires is shaped by multiple interacting factors, including sociocultural, psychological, and design-related influences. These factors are dynamic and depend on context. The proposed typology contributes both theoretically and practically by offering a more integrated way to understand and design for different user needs in digital mental health.

### Limitations and Directions for Future Research

While this study provides valuable insights into men’s engagement with DMH screening questionnaires, several limitations should be noted. The sample included 17 men aged 20–54 and was recruited through purposive and snowball sampling, which may limit transferability beyond this age range and to men in different cultural, social, or economic contexts. The findings may primarily reflect the experiences of participants who were relatively more digitally literate, more educated, or otherwise more inclined to engage with technology than the general population. As a result, the archetypes identified here may partly reflect the perspectives of a technologically more engaged subgroup rather than men more broadly. This does not undermine the findings, just suggests that the barriers and facilitators identified may differ in lower-resource or less digitally mature contexts.

Using self-reported data may also mean that participants did not remember everything accurately or gave answers they saw as more acceptable. In addition, qualitative interviews and thematic analysis are subjective, so the findings are partly shaped by the interaction between interviewer and participant and by the researcher’s interpretation. Finally, data were collected through both face-to-face and Microsoft Teams interviews, and differences in interview mode may have influenced how openly participants responded.

Future research should examine how terminological clarity, clear communication of purpose, and trust-building design features, such as data protection measures and explicit user benefits, influence uptake, engagement, and response validity. It should also systematically evaluate how digital systems can balance early risk detection with the minimization of false positives. In addition, future studies should explore whether DMH screening questionnaires can be tailored to specific subgroups, such as individuals in high-stakes professions, for whom anonymity and data security are critical due to potential career implications. Future research should also investigate whether the archetypes identified in this study can be operationalized in a quantitative questionnaire format, enabling their validation in larger and more diverse samples and assessment of their value for predicting uptake, engagement, and response patterns. Finally, studies should practically test different UX and questionnaire design approaches, including question length, progression cues, feedback modalities, and response formats to determine which designs best support sustained use while minimizing cognitive load.

## Conclusions

This study identified factors influencing men’s willingness to use DMH screening questionnaires for early detection through thematic analysis. Four participant archetypes were distinguished: Skeptic, Self-Manager, Explorer, and Contextual Seeker. These archetypes helped to systematize participants’ attitudes and behavioral patterns. Four key themes emerged: social norms and perceived stigma, anonymity and data protection, user experience and design preferences, and questionnaire structure and content. While these themes align closely with the original research questions (RQ1–RQ3), they were empirically grounded in participants’ accounts, reflecting both their expressed experiences and preferences regarding DMH screening questionnaires.

The main contribution of this study is the development of a behavior-based, context-sensitive typology of men’s engagement with DMH screening solutions that extends beyond demographic segmentation. The findings demonstrate that engagement is not stable or uniform, but shaped by dynamic interactions between trust, perceived risk, usability, and situational context. In particular, institutional trust emerged as a central determinant of willingness to engage, with participants preferring solutions embedded within publicly trusted healthcare systems and supported by clear data protection mechanisms, while high-stakes professionals emphasized the need for full anonymity.

In addition, personalized and actionable feedback with clear next steps was identified as a key factor for sustaining engagement. This suggests that usability alone is not enough, the solution also needs to feel relevant and practically useful to users. Overall, this study provides a more nuanced understanding of men’s engagement with DMH screening questionnaires and offers a framework that can inform the design of more adaptive, trust-sensitive, and user-centered digital mental health solutions.

## Data Availability

The datasets generated and analysed during the current study are not publicly available due to the sensitive nature of the qualitative interview data but are available from the corresponding author on reasonable request.

## Acknowledgments

The authors thank the study participants for their time and valuable contributions. ChatGPT (OpenAI) was used for grammar checking and sentence refinement. To enhance data privacy, chat history and model training were turned off. The tool was not used for data analysis, interpretation, conceptual development, or content generation, and all text was critically reviewed and approved by the author.

## Appendix 1. Semi-Structured Interview Guide English translation version

Thank you for taking the time for this interview. I am Marianne, a master’s student in service design and management at Pärnu College, conducting this study as part of my thesis. I hold a bachelor’s degree in food technology, so I lack medical training, but I am here to understand your experiences, opinions, preferences, and attitudes toward digital mental health early detection solutions, such as mobile apps or web-based questionnaires. Mental health challenges are becoming increasingly common, and research shows that men often face barriers to seeking help. Therefore, it is crucial to capture men’s perspectives to develop solutions that truly meet your needs.

These solutions are often based on scientifically validated and clinically recognized questionnaires that remain paper-based in practice. This study also aims to support the development of a digital prototype for selected questionnaires. This interview is not a test or diagnostic assessment of your mental health; it is simply a conversation to explore how men think about mental health and which design elements would facilitate the use of various digital questionnaires. Please avoid sharing any personal health data.

For accurate analysis, I would like to record this interview. Recordings and transcriptions will be anonymized, your name and all direct identifiers will be replaced with a code (e.g., “Participant 1”). Recordings will be deleted, and results in reports or publications will prevent participant identification. Your data will not be shared with anyone beyond the principal investigator (me). May I record our conversation? Thank you, let’s begin!

| Nr | Theme | Main Question | Follow-up questions | RQ |
| --- | --- | --- | --- | --- |
| 1 | Warm-Up | When you think about regular mental health check-ups, what comes to mind? | What might be useful or harmful in regular monitoring? |  |
| 2 | Social norms and masculinity | In your view, how do societal expectations and perceptions related to masculinity affect men's willingness to seek mental health support online? | For example, how men "should" feel or behave in certain situations. | RQ1 |
| 3 | Social norms and masculinity | Have you ever discussed mental health with friends or family? | (YES) How has talking about mental health with friends or family influenced your willingness to seek help or use digital mental health questionnaires?<br>(NO) If you imagined discussing mental health with friends or family, how might that affect your willingness to seek help or use digital solutions? | RQ1 |
| 4 | Social norms and masculinity | If you decided to seek mental health support, how much would practical factors, such as cost, waiting times, or service availability, influence your choice of a digital mental health solution versus other options? | - | RQ1 |
| 5 | Digital experience (If no, then 5.3) | Have you ever used a digital mental health solution or app? | If yes, which ones? (If no, go to 5.3) | RQ2, RQ3 |
| 5.1 | Digital experience (YES) | Which elements or features helped you with those solutions, and what hindered or disrupted them? | - | RQ2 |
| 5.2 | Digital experience (YES) | Did you continue using the service regularly, or was it a one-time experience? | What influenced that decision? | RQ2 |
| 5.3 | Digital experience (NO) | Imagine these solutions for regular mental health monitoring exist, which situations would you use them in? | What might hinder usage? | RQ1, RQ2 |
| 6 | Motivation and engagement | In what situations might you feel motivated to use a digital mental health questionnaire consistently? | What would facilitate this? | RQ2 |
| 7 | Motivation and engagement | How important are notifications and reminders to you? | Which notification methods would work for you, or currently work in other solutions? | RQ2 |
| 8 | Usability and design | What design elements do you generally like in digital solutions? | - | RQ2 |
| 9 | Usability and design | What design elements would make a digital mental health solution more appealing? | - | RQ2 |
| 10 | Usability and design | When completing a digital questionnaire about your well-being or mental health, what guidance or explanations do you expect? | Preferably before starting or during completion? When is guidance unnecessary? | RQ3 |
| 11 | Usability and design | In what ways would visual elements (scales/icons) help or hinder usage? | - | RQ3 |
| 12 | Usability and design | Would you prefer answering questions in a standard questionnaire format or through a conversational interface, like a chatbot? | Why would that format be more comfortable for you? | RQ3 |
| 13 | Usability and design | What makes the content of a digital solution relatable or comfortable for men? | Consider language (e.g., words, tone, expressions), visuals (e.g., colors, fonts, images), and presentation (e.g., length, format). Anything that feels uncomfortable or off-putting? | RQ2 |
| 14 | Usability and design | What kind of feedback would you prefer on your results? | - | RQ2 |
| 15 | Usability and design | Which results presentation format would be most understandable: text, visuals, chatbot, animation, video, or a combination? | Why that format? | RQ3 |
| 16 | Motivation and engagement | Would it help if the provided guidance on next steps or coping strategies? | What advice or guidance would be useful to you? | RQ2 |
| 17 | Usability and privacy | When thinking about completing a digital mental health questionnaire, what would make you prefer fully anonymous use versus a personalized experience? | - | RQ1 |
| 18 | Usability and privacy | How do you feel about your responses and results being saved for later review or tracking changes over time? | - | RQ1, RQ2 |
| 19 | Usability and platform | Is it important which digital platform the questionnaire is on? | Do you have any suggestions for health- or mental health-related websites or digital platforms? (Examples provided if needed: Terviseportaal, peaasi.ee, perearst24, e-consultation services, or e-family doctor systems. Participants were also asked about prior use and preferences.) | RQ1 |
| 20 | Usability and design | When using various digital questionnaires, at what point and why does your interest or focus wane? | Related to time investment, content volume, question style, or something else? | RQ3 |
| 21 | Motivation and engagement | What would help you complete a digital mental health questionnaire once started? | - | RQ3 |
| Provide questionnaires (EEK-2, PHQ-2, PHQ-9, WHO-5) for review |  |  |  |  |
| 22 | Questionnaire | Which provided questionnaire would you prefer for real-life mental health monitoring? | Why that choice? | RQ3 |
| 23 | Questionnaire | Do you prefer direct or descriptive questions? | Examples if needed. | RQ3 |
| 24 | Questionnaire | Which questions feel uncomfortable or ones you wouldn't want to answer? | How could they be phrased better from your perspective? | RQ1 |
| 25 | Conclusion | Is there anything else developers should know when creating digital mental health questionnaires for men? | Anything we haven't discussed that you'd like to add or emphasize? | – |

Thank you very much for sharing your experiences and thoughts today. Your input is truly valuable and will help create digital mental health questionnaires that are more useful, engaging, and supportive for men. Just a reminder, everything shared remains confidential. If more thoughts arise after the interview, feel free to contact me. I greatly appreciate your time and contribution.

## Appendix 2. Barriers and Facilitators to Help-Seeking by Participant Archetype

| Aspect | Skeptic |  | Self-Manager |  | Explorer |  | Situational Seeker |  |
| --- | --- | --- | --- | --- | --- | --- | --- | --- |
| **Data Privacy** | × Data misuse fear | ✓ Transparency<br>✓ User control | × Low transparency<br>× Perceived data risk | ✓ Full user data control<br>✓ Personal tracking storage | × Unclear data use | ✓ Transparency<br>✓ User control | × Fear of data misuse | ✓ Anonymous access<br>✓ Trace-free use |
| **Institutional legitimacy** | × Standalone platforms<br>× Distrust of general practitioners | ✓ Formal channels<br>✓ Institutional affiliation<br>✓ In-person contact | × Stand-alone digital platforms | ✓ Expert support need<br>✓ Healthcare integration | × System distrust (data)<br>× Informal help preference | ✓ Familiar platform<br>✓ Institutional backing<br>✓ Flexible engagement | × Informal support<br>× Stand-alone platform | ✓ Familiar platform<br>✓ Expert backup<br>✓ Healthcare integration |
| **UX/design** | × Too many steps<br>× Information overload<br>× Unclear timelines | ✓ Low visual noise<br>✓ One task at a time<br>✓ Simple language | × High cognitive load<br>× Low perceived meaning<br>× Technical friction | ✓ Mobile friendly<br>✓ Structured flows<br>✓ Visible progress<br>✓ Evidence-based feedback | × Text-heavy<br>× Unclear purpose<br>× Long and generic questions<br>× Many complex steps | ✓ Mobile friendly<br>✓ Visuals<br>✓ Personal feedback<br>✓ Interactive formats<br>✓ Progress indicator | × Text-heavy<br>× Too long tool<br>× Poor clarity | ✓ Short (5-10 minutes)<br>✓ Clear structure<br>✓ 1 question at a time<br>✓ Clean visual |
| **Accessibility** | × High cost<br>× Long waits | ✓ Easy, anonymous access<br>✓ Cost-free | × Price sensitive<br>× Low time availability | ✓ Everyday integration<br>✓ Flexible access<br>✓ Low cost | × Low awareness<br>× Time burden<br>× Paywall | ✓ Optional anonymity<br>✓ Quick use<br>✓ No queues | × Long waits<br>× High effort access<br>× Paywall | ✓ Cost-free<br>✓ Mobile access<br>✓ Immediate availability |
| **Help-Seeking orientation** | × Stigma<br>× Help distrust<br>× Self-reliance | ✓ Safe sharing space<br>✓ Proven effectiveness | × Self-reliance<br>× Crisis-driven<br>× Poor self-state<br>× Low value | ✓ Self-monitoring<br>✓ Goal-driven<br>✓ Openness to support | × Only when problem arise<br>× Delay in seeking help | ✓ Perceived usefulness<br>✓ Situational openness | × Stigma<br>× Crisis only<br>× Friends first<br>× Mental health de-prioritization | ✓ Perceived urgency |

